# Tracking and Classifying Global COVID-19 Cases by using 1D Deep Convolution Neural Networks

**DOI:** 10.1101/2020.06.09.20126565

**Authors:** Mark Amo-Boateng

## Abstract

The novel coronavirus disease (COVID-19) and pandemic has taken the world by surprise and simultaneously challenged the health infrastructure of every country. Governments have resorted to draconian measures to contain the spread of the disease despite its devastating effect on their economies and education. Tracking the novel coronavirus 2019 disease remains vital as it influences the executive decisions needed to tighten or ease restrictions meant to curb the pandemic. One-Dimensional (1D) Convolution Neural Networks (CNN) have been used classify and predict several time-series and sequence data. Here 1D-CNN is applied to the time-series data of confirmed COVID-19 cases for all reporting countries and territories. The model performance was 90.5% accurate. The model was used to develop an automated AI tracker web app (AI Country Monitor) and is hosted on https://aicountrymonitor.org. This article also presents a novel concept of pandemic response curves based on cumulative confirmed cases that can be use to classify the stage of a country or reporting territory. It is our firm believe that this Artificial Intelligence COVID-19 tracker can be extended to other domains such as the monitoring/tracking of Sustainable Development Goals (SDGs) in addition to monitoring and tracking pandemics.

## Introduction

The COVID-19 disease was declared as a global pandemic^1–4^ leading to the imposition of restrictions in almost all countries and states worldwide. Since then, confirmed COVID-19 cases, as well as, reported recoveries and deaths have been tracked and monitored on several apps and platforms^1,2,5–7^. These tracking dashboards and apps provide data that can be used to estimate the rate of spread and other statistics which influences the top-level decisions by governments which may lead to tightening or easing of these state-wide restrictions and policies^6,8–10^.

The existing COVID-19 trackers^2,6,11,12^ focus on presenting the COVID-19 case data as reported by countries and territories only as dashboards and charts; they and do not perform any predictions or classifications. We also note that no studies or existing dashboards/trackers employ the use of artificial intelligence to predict the state of COVID-19 for reporting countries and territories. The method proposed in this study involves data preprocessing, feature extraction, and predictions using one dimensional deep convolution neural networks. Even though other algorithms exist that can be used in performing feature extraction and classification of sequence or time-series data, convolution neural networks have been found to be superior^13–16^ in extracting unique features and patterns in sequences^17–19^, images and video data. In fact, deep convolution neural networks algorithms are behind some of the state-of-the-art detection and tracking algorithms found in everyday images and videos. Here, 1D-CNN is used to extract unique feature identifiers that help predict the true state of countries and territories with respect to the novel coronavirus disease, uncovering hidden and underlying patterns that might otherwise not be readily noticed.

We propose an artificial intelligence prediction algorithm for the novel coronavirus 2019 stages for countries and reporting territories based on 1-D deep convolution neural networks trained on confirmed COVID-19 cases. We also propose the concept of pandemic response curves also based on confirmed COVID-19 cases that provides further insights into the state of each reporting country or territory. Our main aim is to provide real-time predictions of the stages of COVID-19 that is unbiased or subjective to human prejudices for all countries and reporting territories.

## Results

### Country and State Predictions

The COVID-19 state for each country and reporting territories on 11th, 18th, 25th and 31st May as predicted by the AI developed in this paper is given in Figure 1. Detailed predictions for 11th May, 2020 is shown in Table 1.For the diagram, Red: Stage 1 - Emerging (cases actively rising); Amber: Stage 2 - Controlling (cases passed peak and actively declining); Green: Stage 3 - Winning (cases consistently zero or near zero after stage 2). Weekly changes in the stages of each country is also shown in Figure 2. It can be seen that very few countries are changing their stages in the COVID-19 pandemic each week. Detailed country COVID-19 stages and changes is given in Table 2. Overall, only 54 countries and reporting states changed their COVID-19 status.

**Table 1.**
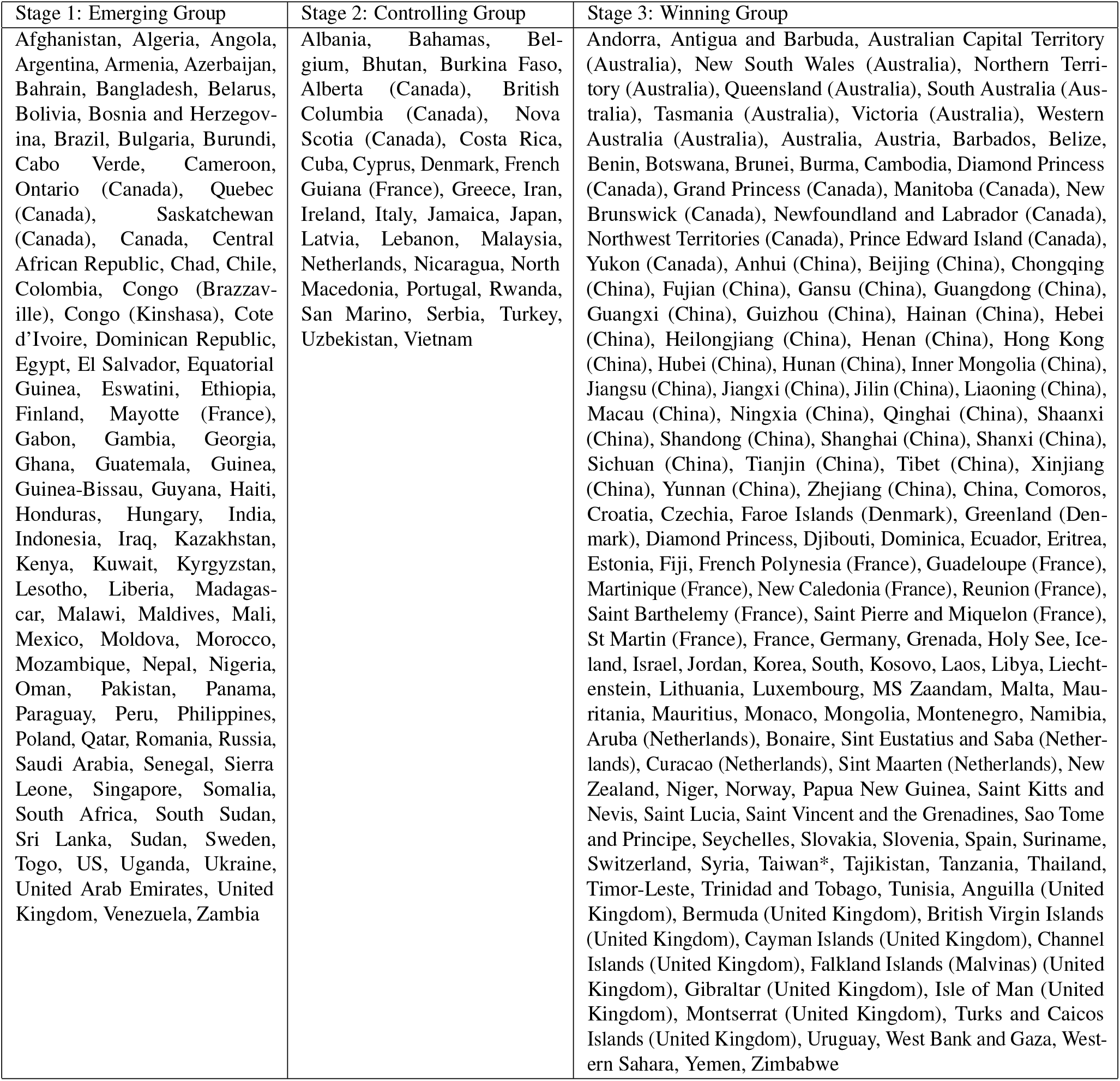
Prediction of COVID-19 status for countries and reporting territories for 11th May, 2020.

**Table 2.** Countries and reporting territories that have changed their COVID-19 state in May.

| SN | Country | ISO(A3) | 11-May | 18-May | 25-May | 31-May |
| --- | --- | --- | --- | --- | --- | --- |
| 1 | Albania | ALB | Controlling | Controlling | Controlling | Emerging |
| 19 | Bahamas | BHS | Controlling | Winning | Controlling | Controlling |
| 26 | Benin | BEN | Winning | Emerging | Winning | Winning |
| 27 | Bhutan | BTN | Controlling | Emerging | Emerging | Emerging |
| 29 | Bosnia and Herzegovina | BIH | Emerging | Controlling | Controlling | Controlling |
| 30 | Botswana | BWA | Winning | Winning | Emerging | Emerging |
| 33 | Bulgaria | BGR | Emerging | Emerging | Emerging | Controlling |
| 35 | Burma | MMR | Winning | Winning | Winning | Controlling |
| 48 | Nova Scotia (Canada) |  | Controlling | Controlling | Winning | Winning |
| 51 | Quebec (Canada) |  | Emerging | Emerging | Emerging | Controlling |
| 52 | Saskatchewan (Canada) |  | Emerging | Controlling | Controlling | Winning |
| 54 | Canada | CAN | Emerging | Emerging | Emerging | Controlling |
| 96 | Costa Rica | CRI | Controlling | Controlling | Controlling | Emerging |
| 100 | Cyprus | CYP | Controlling | Winning | Winning | Winning |
| 101 | Czechia | CZE | Winning | Controlling | Controlling | Controlling |
| 106 | Djibouti | DJI | Winning | Winning | Emerging | Emerging |
| 109 | Ecuador | ECU | Winning | Winning | Controlling | Controlling |
| 118 | Finland | FIN | Emerging | Controlling | Controlling | Controlling |
| 119 | French Guiana (France) | GUF | Controlling | Emerging | Emerging | Emerging |
| 131 | Gambia | GMB | Emerging | Emerging | Winning | Winning |
| 132 | Georgia | GEO | Emerging | Controlling | Controlling | Controlling |
| 139 | Guinea-Bissau | GNB | Emerging | Emerging | Winning | Winning |
| 144 | Hungary | HUN | Emerging | Controlling | Controlling | Controlling |
| 148 | Iran | IRN | Controlling | Emerging | Emerging | Emerging |
| 153 | Jamaica | JAM | Controlling | Winning | Controlling | Controlling |
| 155 | Jordan | JOR | Winning | Emerging | Emerging | Controlling |
| 159 | Kosovo | XKS | Winning | Controlling | Controlling | Controlling |
| 164 | Lebanon | LBN | Controlling | Emerging | Emerging | Emerging |
| 165 | Lesotho | LSO | Emerging | Winning | Winning | Winning |
| 167 | Libya | LBY | Winning | Winning | Winning | Emerging |
| 177 | Malta | MLT | Winning | Controlling | Controlling | Controlling |
| 178 | Mauritania | MRT | Winning | Winning | Winning | Emerging |
| 183 | Mongolia | MNG | Winning | Emerging | Emerging | Winning |
| 185 | Morocco | MAR | Emerging | Emerging | Controlling | Controlling |
| 187 | Namibia | NAM | Winning | Winning | Winning | Emerging |
| 195 | Nicaragua | NIC | Controlling | Emerging | Winning | Winning |
| 198 | North Macedonia | MKD | Controlling | Controlling | Controlling | Emerging |
| 204 | Paraguay | PRY | Emerging | Emerging | Controlling | Controlling |
| 210 | Romania | ROU | Emerging | Emerging | Controlling | Controlling |
| 212 | Rwanda | RWA | Controlling | Winning | Controlling | Controlling |
| 215 | Saint Vincent and the Grenadines | VCT | Winning | Winning | Winning | Emerging |
| 217 | Sao Tome and Principe | STP | Winning | Winning | Winning | Emerging |
| 232 | Suriname | SUR | Winning | Winning | Winning | Emerging |
| 235 | Syria | SYR | Winning | Emerging | Emerging | Emerging |
| 237 | Tajikistan | TJK | Winning | Emerging | Emerging | Emerging |
| 250 | Bermuda (United Kingdom) | BMU | Winning | Winning | Winning | Controlling |
| 252 | Cayman Islands (United Kingdom) | CYM | Winning | Controlling | Emerging | Emerging |
| 259 | United Kingdom | GBR | Emerging | Emerging | Controlling | Controlling |
| 260 | Uruguay | URY | Winning | Winning | Controlling | Controlling |
| 261 | Uzbekistan | UZB | Controlling | Emerging | Emerging | Emerging |
| 263 | Vietnam | VNM | Controlling | Controlling | Winning | Winning |
| 264 | West Bank and Gaza | PSE | Winning | Winning | Controlling | Controlling |
| 266 | Yemen | YEM | Winning | Emerging | Emerging | Emerging |
| 267 | Zambia | ZMB | Emerging | Emerging | Emerging | Winning |
| 268 | Zimbabwe | ZWE | Winning | Controlling | Emerging | Emerging |

**Figure 1.**
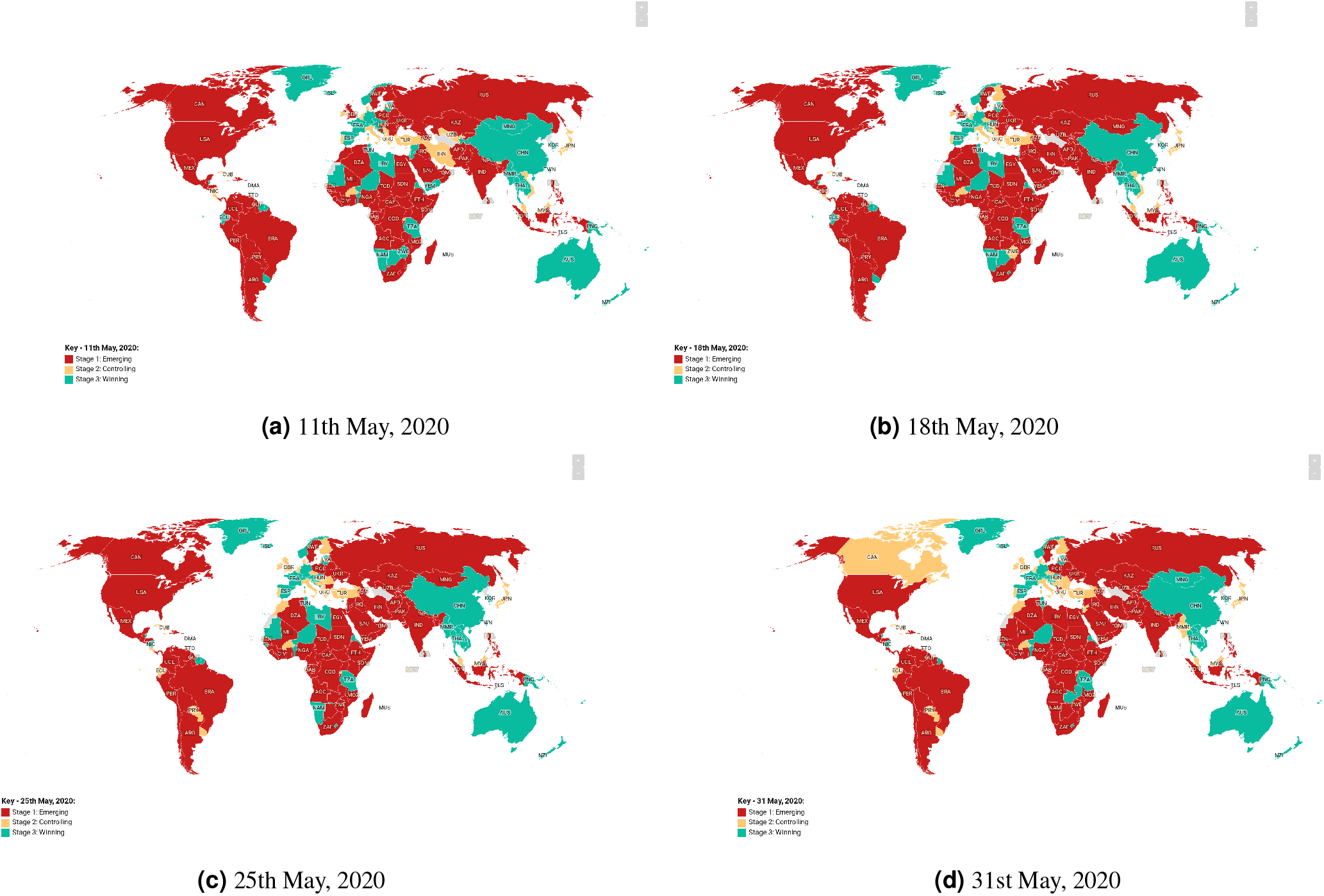
Weekly COVID-19 Global Predictions of Countries and Territories for May, 2020)

**Figure 2.**
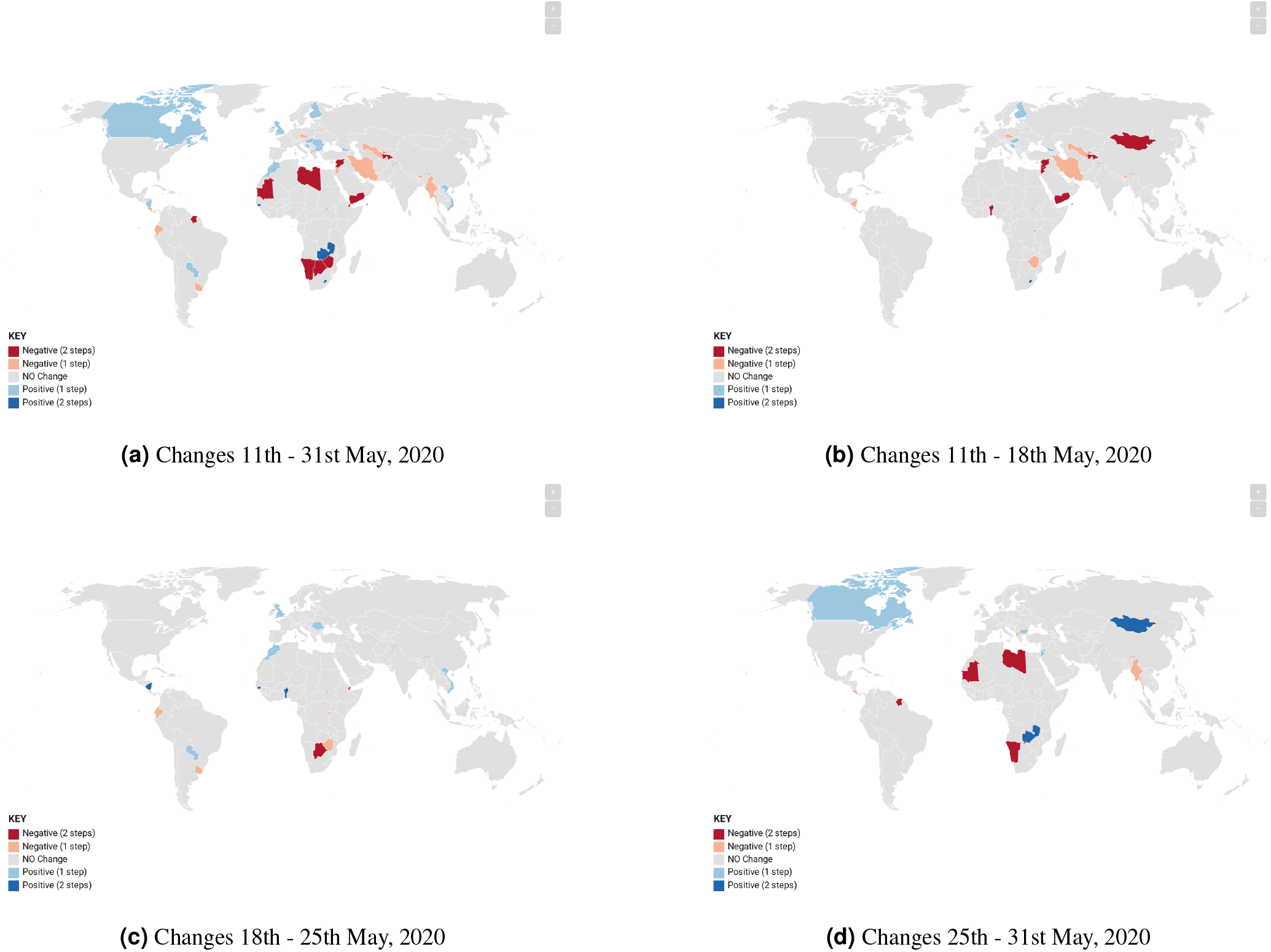
Global Changes in COVID-19 Stages for Countries and Territories in May, 2020.

Also, COVID-19 state for each county and territories in the USA on on 11th, 18th, 25th and 31st May as classified by the AI used in this paper is given in Figure 3. The color code for the diagram is: Red: Stage 1 - Emerging (cases actively rising); Amber: Stage 2 - Controlling (cases passed peak and actively declining); Green: Stage 3 - Winning (cases consistently zero or near zero after stage 2). Changes in the US counties and territories as seen each week is shown in Figure 4. It is very evident that though some counties did not have any changes over the month of May, many counties had a negative change, moving from Stage 3: Winning to Stage 1: Emerging; while others moved in the opposite direction.

**Figure 3.**
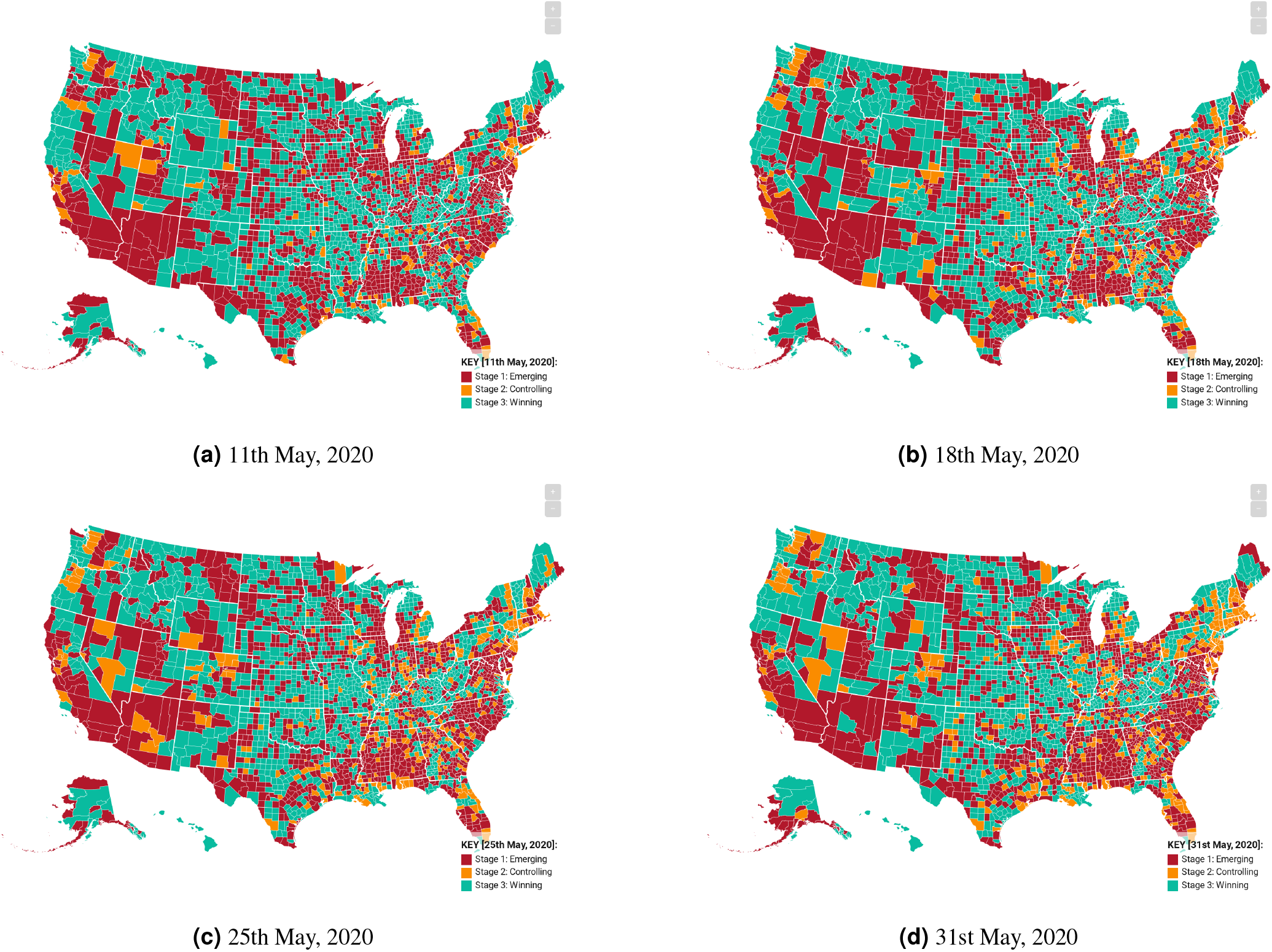
Weekly COVID-19 USA Predictions of Counties and Territories for May, 2020.

**Figure 4.**
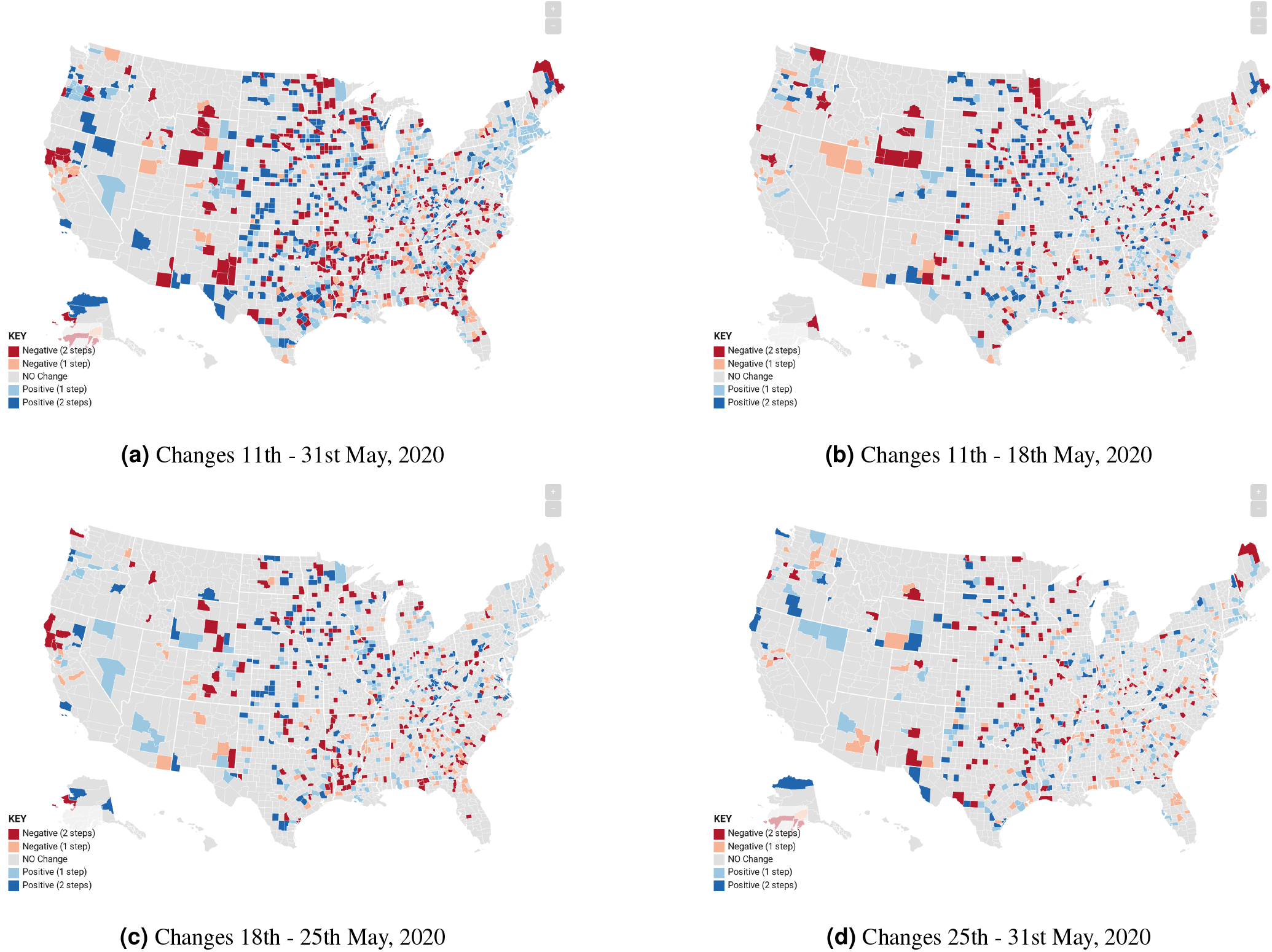
Changes in Weekly COVID-19 Stages in USA Counties and Territories for May, 2020.

### Pandemic Response Curves

The Pandemic Response Curves, as shown in Figure 5, provides a visual inspection of the similarities among countries in the various stages of the COVID-19 pandemic. The curve highlights patterns that may have otherwise not been noticeable and helps clarify unbiasedly where each country or reporting territory is at in the COVID-19 fight. Figure 6 provides some of the PRC for selected countries in each of the stages. Also, Figure 7 shows the country Ghana PRC compared to other countries’ PRC which recorded their first cases of COVID-19 the same day.

**Figure 5.**
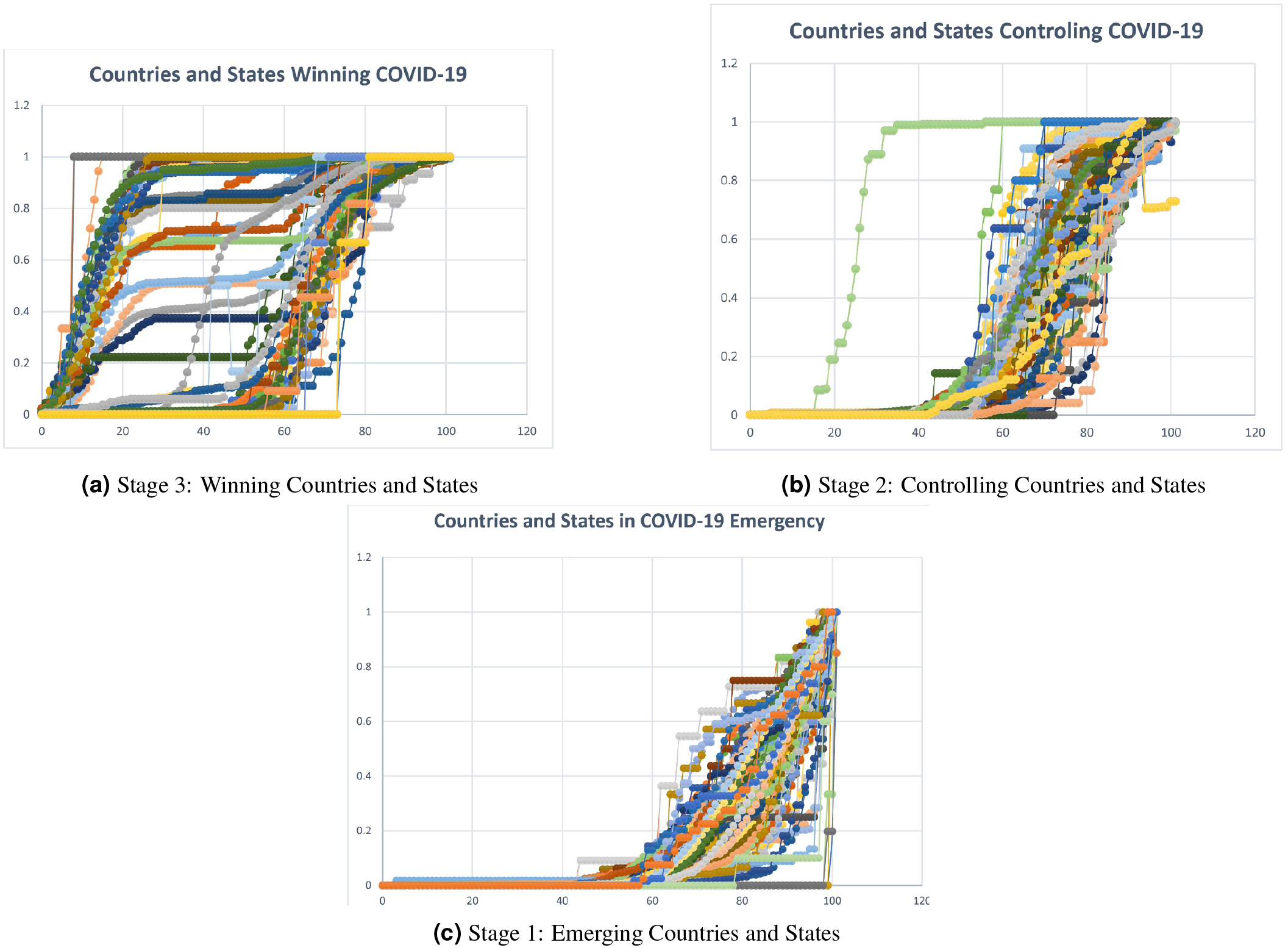
General COVID-19 Pandemic Response Curves for Countries and Territories (11th May, 2020)

**Figure 6.**
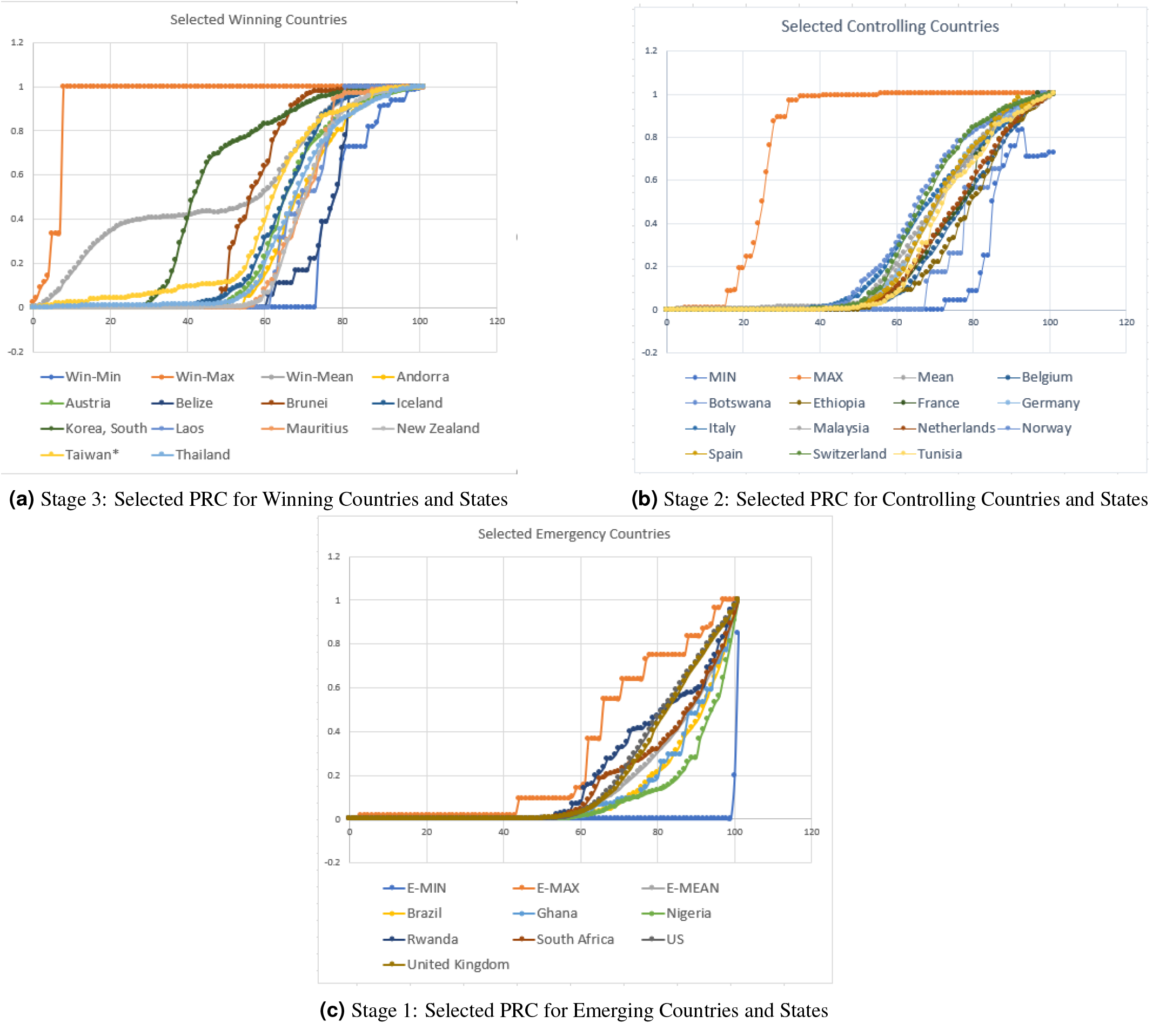
Sample COVID-19 Pandemic Response Curves for Selected Countries and Territories (4th May, 2020)

**Figure 7.**
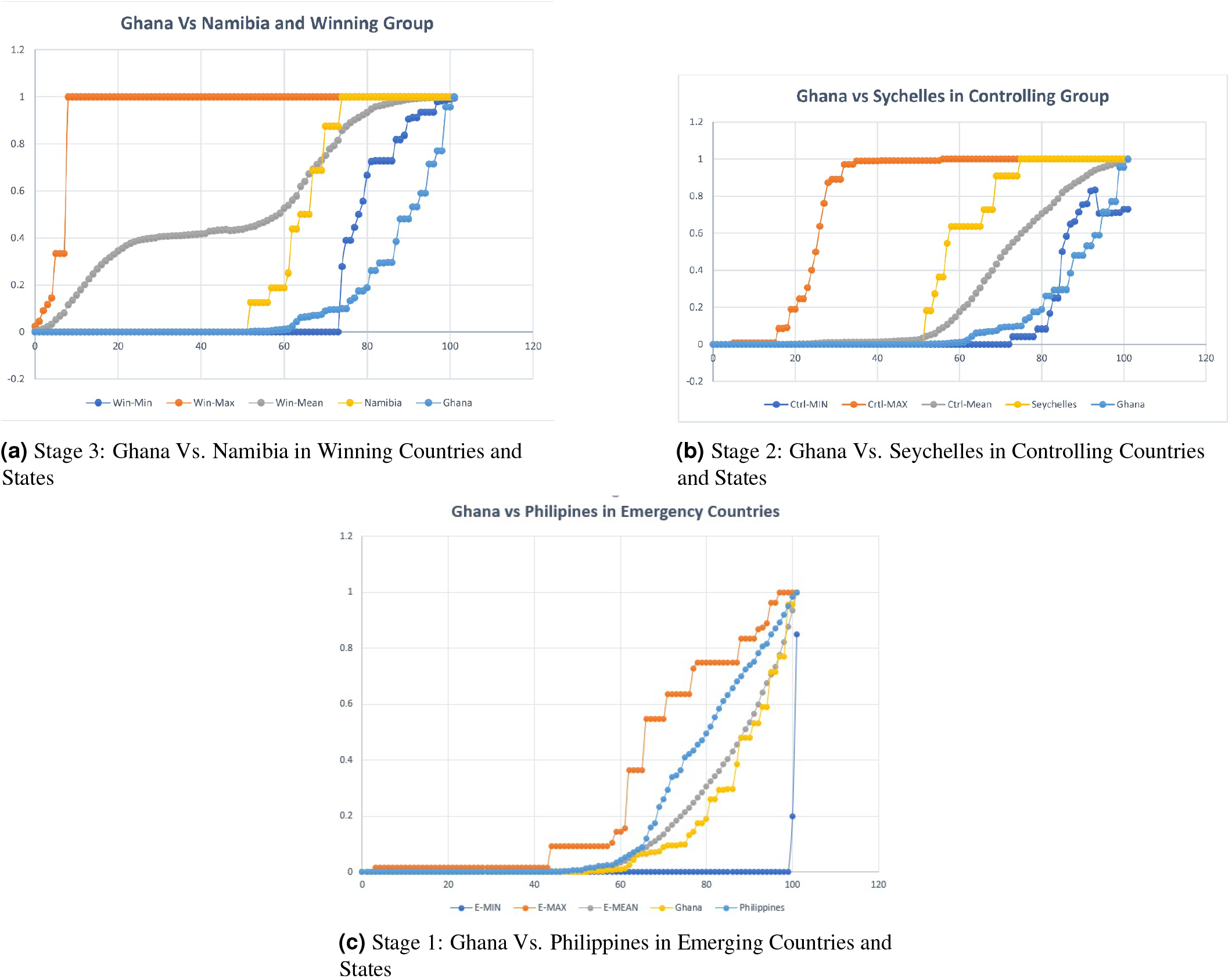
Ghana versus selected countries COVID-19 Pandemic Response Curves (11th May, 2020)

**Figure 8.**
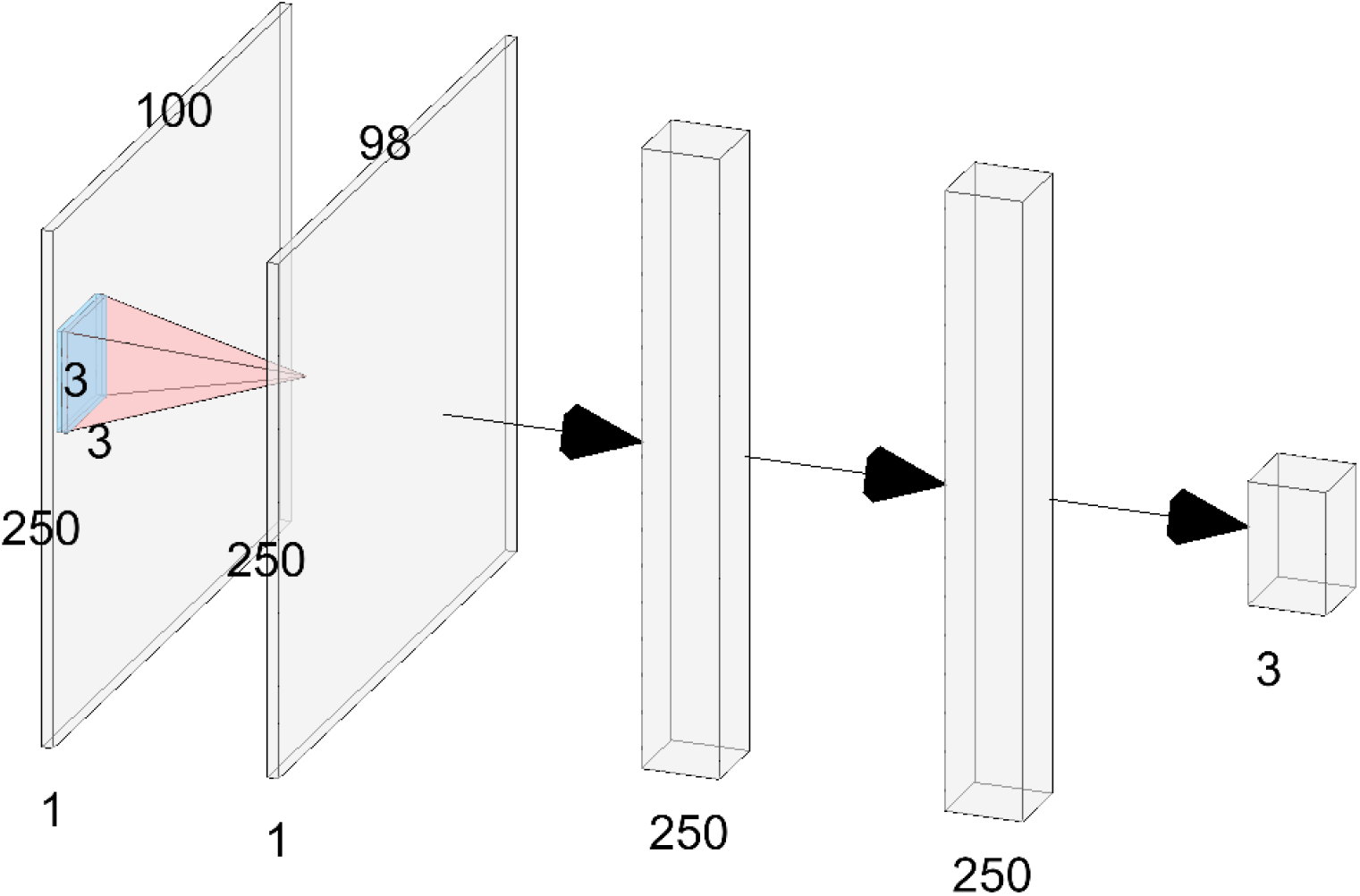
COVID-19 AI Network Architecture.

**Figure 9.**
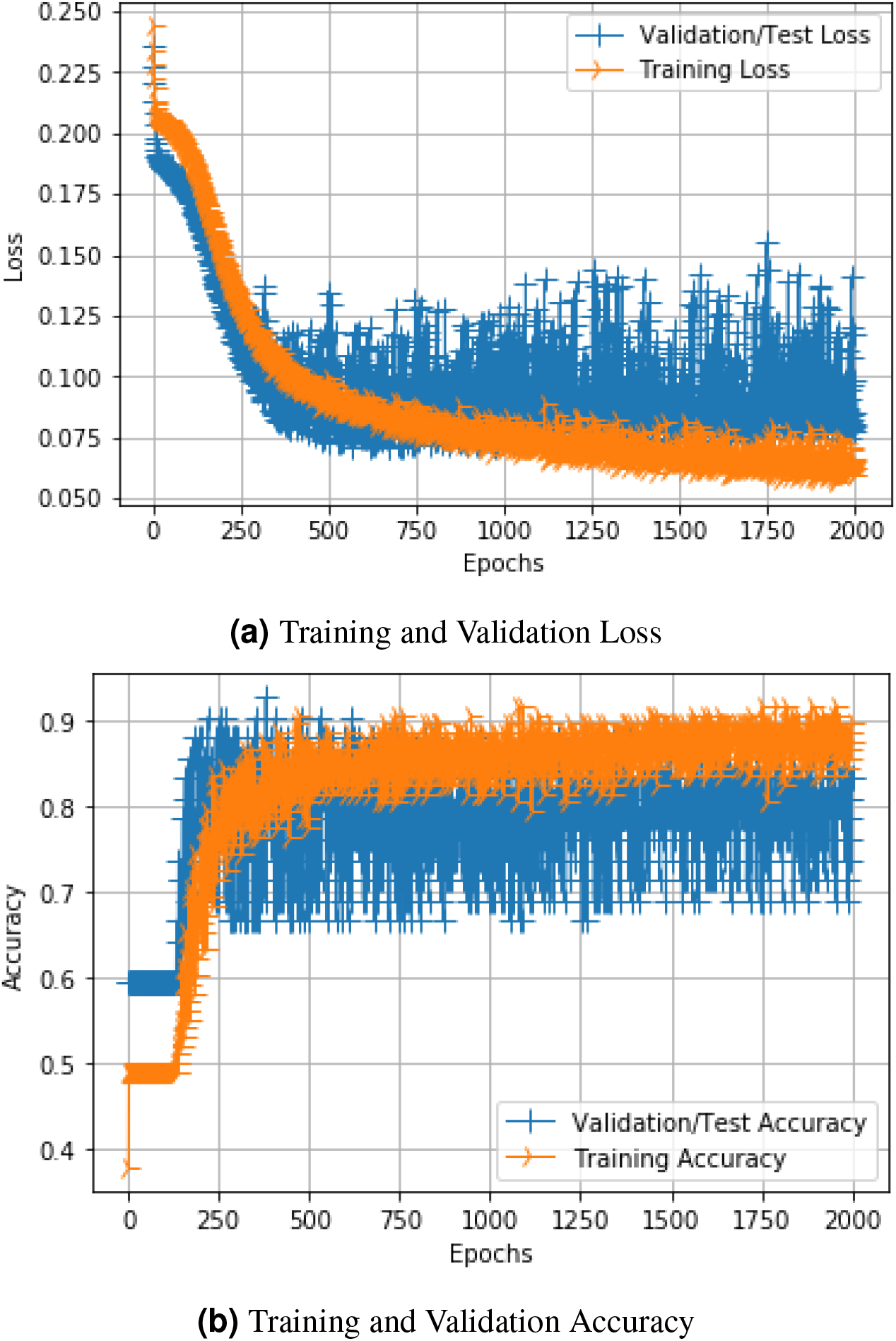
COVID-19 AI Model Training Loss and Accuracy.

## Discussion

The COVID-19 pandemic is still ravaging many countries and territories, and has led to strict movement restrictions including border closures and curfews in many countries. Public gatherings, schools, restaurants and hotels have also been closed as a results on its unprecedented method of spreading. To help contain the virus, many apps have been developed that track infections across the globe. Notable amongst them include the John Hopkins CSSE COVID-19 dashboard which has served as a reference database. To understand further understand hidden insights to all reported cases an artificial intelligence was developed in this paper. Artificial intelligence is already being applied COVID-19 diagnosis of Chest XRays and CT-Scans^20,21^. In this paper, we applied AI to the cumulative confirmed coronavirus cases to uncover hidden insights and tell the status of each country for the month of May.

This begun by preprocessing the data and manually classifying some of the data from the countries (see Table 3. The data was meant to help the AI be a generalizer of the patterns in the confirmed coronavirus cases. Further, the AI was developed using Convolution Neural Networks which has been demonstrated to be superior in finding hidden patters in images and videos. By apply a 1D-CNN, we had hoped for a very efficient pattern finding of the confirmed COVID-19 cases, and used that to classify the cases into three main groups: Stage 1: Emerging (active rising cases); Stage 2: Controlling (passed peak and active decline cases); Stage 3: Winning (consistently zero or near zero cases after active decline).

**Table 3.** Pre-classifications used for training the COVID-19 AI using data based on 4th May, 2020.

| Stage 1: Emerging Group | Stage 2: Controlling Group | Stage 3: Winning Group |
| --- | --- | --- |
| Argentina, Azerbaijan, Bahrain, Bangladesh, Belarus, Brazil, Bulgaria, Central African Republic, China - Anhui, Democratic Republic of Congo, El Salvador, Equatorial Guinea, France - French Polynesia, Islamic Republic of Iran, Iraq, Ireland, Israel, Monaco, Mongolia, Norway, Papua New Guinea, Paraguay, Poland, Portugal, Qatar, Russian Federation, Rwanda, Saint Lucia, Slovenia, Spain, Taiwan, United Kingdom - Bermuda, United States of America, R. B. de Venezuela, Canada - Yukon | Albania, Belgium, Bosnia and Herzegovina, Burkina Faso, Croatia, Denmark - Faroe Islands, Greenland (Den.), Dominican Republic, The Gambia, Greece, India, Italy, Jamaica, Japan, Kazakhstan, Republic of Korea, Lebanon, Malta, Nicaragua, North Macedonia, Oman, Pakistan, Romania, Senegal, Singapore, Sudan, Suriname, Tanzania, Ukraine, Uzbekistan, Vietnam | Andorra, Australia - Australian Capital Territory, Australia - New South Wales, Australia - Northern Territory, Australia - Queensland, Australia - South Australia, Australia - Tasmania, Australia - Victoria, Australia - Western Australia, Austria, Bhutan, Cambodia, China - Beijing, China - Chongqing, China - Fujian, China - Gansu, China - Guangdong, China - Guangxi, China - Guizhou, China - Hainan, China - Hebei, China - Heilongjiang, China - Henan, Hong Kong (SAR), China - Hubei, China - Hunan, China - Inner Mongolia, China - Jiangsu, China - Jiangxi, China - Jilin, China - Liaoning, Macau (SAR), China - Ningxia, China - Qinghai, China - Shaanxi, China - Shandong, China - Shanghai, China - Shanxi, China - Sichuan, China - Tianjin, China - Tibet, China - Xinjiang, China - Yunnan, China - Zhejiang, Colombia, Congo, Cuba, Czech Republic, Ecuador, Ethiopia, Guinea, Indonesia, Jordan, Kenya, Kyrgyz Republic, Liberia, Liechtenstein, Luxembourg, Madagascar, Malaysia, Mauritius, Moldova, Montenegro, Namibia, Niger, Somalia, South Africa, Thailand, Trinidad and Tobago, Uganda, Zimbabwe, United Kingdom - British Virgin Islands |

Training the AI on the pre-classified data using the last 100 days yield a 90.5% accuracy. The trained model was then applied to the whole data for all countries and reporting territories, as well as US counties and territories. The results of this operation for each country is shown in Figure 1 and Figure 3 for US counties and territories. In addition, tracking the changes overtime is paramount to helping us understand which countries or states are performing well or otherwise, as such, changes in the performance of all countries and states were also carried out. These results for the month of May can be seen in Figure 2 and Table 2 for all countries and reporting territories. Similar results for US counties and territories are shown in Figure 4. From the above results, it can be clearly seen that very few countries, states and US counties are changing their COVID-19 status. There are even instances where some countries or states went from consistently very low or zero cases to actively rising cases (e.g. Djibouti, Namibia, Zimbabwe).

The AI developed in this paper currently relies on patterns within the cumulative confirmed coronvavirus cases for all reporting countries and territories. A further investigation into the performance of the AI led to the development of the Pandemic Response Curves (PRC). The PRC showed that countries with similar COVID-19 status had identical curves. This suggests that the generalized PRC shapes can be used to classify the status of all countries in an unbiased way. The PRCs for all countries, highlighting the shapes and similarities of curves (irrespective of population and other metrics) are shown in 5. Selected countries with kwown COVID-19 status are shown in 6. To further highlight the uniqueness of the PRCs, Ghana (a Stage 1: Emerging country) was compared to other countries that recorded their first case on the same day. This result is shown in 7 for Namibia (Stage 3: Winning - see Figure 7a), Seychelles (Stage 2: Controlling - see Figure 7b), and Phillipines (also Stage 1: Emerging - see Figure 7c). It is hoped that the PRCs be developed further to help automatically classify pandemics.

## Methods

### Dataset

The dataset used for this project is the one compiled daily by the John Hopkins Center for Systems Science and Engineering (CSSE) and made available to the public via [GitHub](https://github.com/CSSEGISandData/COVID-19). Data sources used in compiling the CSSE database include: WHO, CDC, ECDC, NHC, DXY, 1point3acres, Worldometers.info, the COVID Tracking Project (testing and hospitalizations), and city, county, state and national public health departments. The time series data consist of daily cumulative cases of COVID-19 from all countries and territories. In addition, data for the United States counties are presented. The time series data begins on January 22nd, 2020 till date.

### Preprocessing

Each country or reporting territory data is normalized with respect to its maximum cumulative cases. The resulting time-series data is in the range of 0 to 1. To account for the inconsistencies in the data reporting intervals across all countries, the normalized data time-series is then smoothed with time-windowed simple moving average. The following time-windows were considered: 3, 5, 7, 10, 12, 14 day moving averages. Data Labeling: countries and reporting territories that have been found to have been reported in the media as having reached one of the 3 stages of COVID-19 were manually identified, verified (visually and using third-party websites like EndCoronavirus.org and tagged with one of the labels: Stage 1 - Cases actively rising; Stage 2 - Cases have passed peak and begun to actively decline; Stage 3 - Cases consistently zero or near zero after active declining. These stages were labeled as Stage 1: Emerging, Stage 2: Controlling, Stage 3: Winning.In the most recent version of the model update 134 countries were manually labelled for AI training and validation. The labelled data was one-hot encoded for the 3 classes; and then split into 0.7:0.3 ratio for training and validation. Pre-classified countries and territories used in the training of the AI model is shown in Table 3.

### 1D-CNN COVID-19 AI Model

1D-CNN was used in this AI model with an initial input sequence length of 100; thus using data of the last 100 days for training, validation and prediction. This layer also had 64 filters, a kernel size of 3 ⨯ 3, and ReLu activation unit. This was followed by another 1D-CNN also with 64 filters, kernel size of 3 ⨯ 3, padding, strides of 1, and ReLu activation unit. A GlobalMaxPooling1D layer was used to flatten the 1D-CNN and the output fed into a 256 Dense layer. A 20% Dropout layer was then added with ReLu activation unit and then fed to another 128 Dense layer then to the 3 classification output layers using the Sigmoid activation function.

The model was compiled using the Adam as the optimizer(initial learning rate of 0.001), Mean Squared Error (MSE) as the loss function, and Accuracy as the metrics. In addition, four callbacks were also setup and used with model during training. They were: ModelCheckpoint (saved the best models during training), EarlyStopping (terminateg training if no improvement is being made), ReduceLROnPlateau (adjusts the learning rate if optimizer appears stuck in a local minima), and Tensorboard (live monitoring of training progress). The model was then repeatedly trained over 2000 epochs and the best performing models selected for use in the AI Covid Tracker.

## Data Availability

Data and results used for study is available via GitHub page

https://github.com/markamo/aicountrymonitor

https://github.com/CSSEGISandData/COVID-19/tree/master/csse_covid_19_data

## Acknowledgements

I will like to thank the John Hopkins Center for Systems Science and Engineering (CSSE) for making COVID-19 data openly available on GitHub.

## Author contributions statement

A.B. conceived the experiment(s), conducted the experiment(s), and analysed the results. All authors reviewed the manuscript.

## Additional information

We declare no competing interests

## Notes

### Competing Interest Statement

The authors have declared no competing interest.

### Funding Statement

Neither the authors or their institutions received funding for this work.

### Author Declarations

This work does not need any approval of the IRB/oversight body.

